# What Week 8 Knows: Forecasting Six-Month GLP-1 Outcomes

**DOI:** 10.64898/2026.07.14.26357506

**Authors:** Brian Erly, Shanmugesh Raja

**Author notes:** Corresponding author: Brian Erly, MD, MPH —.

## Abstract

**Background:** Patients on GLP-1 medications lose very different amounts of weight, and most published prediction models include only patients who complete six months. That design omits everyone who disengages earlier, which is the majority of the cohort. We built a tool that includes patients who disengage and delivers useful predictions at the week-8 visit, where the clinical decision is actually made.

**Methods:** Beginning with 237,800 adults enrolled in a US telehealth GLP-1 program, we required a documented week-8 weight, a refill-confirmed dose, and reported ethnicity, yielding an analytic cohort of 22,538. We answered three questions: the patient’s likely six-month weight loss and our confidence in it; the probability of dropout before six months; and when weight loss plateaus. For the first, we fit a cubic in week-8 percent loss plus 16 covariates, with quantile-regression bands at the 10th and 90th percentiles for the prediction interval, checking fractional-logit and isotonic recalibration as alternatives. For the second, we fit a logistic regression and compared it to gradient boosting. For the third, we fit a per-patient exponential trajectory among patients with at least four weight observations. We trained on enrollments before 2024-07-01 and tested on later ones, compared completer outcomes to published RCTs, and tested the week-8 anchor (week 8 is the anchor itself) against measurements at weeks 2, 4, 6, 8, 10, 12, 16, and 20.

**Results:** Mean six-month weight loss in completers was 11.7% on semaglutide and 14.1% on tirzepatide, in line with STEP-1 and SURMOUNT-1.^1,2^ Six-month disengagement was 66%. The prediction model reached test R^2^ = 0.65 with a mean absolute error of 2.76 percentage points. Calibration was strong: calibration-in-the-large was *−*0.52 pp and the calibration slope was 0.96. The 80% quantile-regression interval covered 76% of test patients; the 95% interval covered 93%. The disengagement model reached test AUC 0.79, against 0.74 for gradient boosting. Median plateau time among engaged patients was 387 days, longer in lower-BMI tertiles. The week-8 anchor gave R^2^ = 0.65, compared to 0.48 to 0.61 at earlier weeks and 0.67 to 0.91 at later weeks. We chose week 8 because 80% of slow responders reach their post-titration decision point at or before that visit. Two of twenty subgroup cells had reduced predictive accuracy; two more were too sparse to validate.

**Conclusions:** Observed week-8 weight loss is the strongest predictor of six-month outcome. The model’s accuracy (R^2^ = 0.65, MAE 2.76 pp) is appropriate for calibrating expectations and identifying patients for the post-titration decision, but not precise enough to drive that decision on its own. Disengagement is predictable at week 8 with AUC 0.79. Engaged patients plateau at a median of 387 days. Week 8 is the earliest visit at which titration is mostly complete, accuracy is in a useful range, and the post-titration decision remains actionable; later anchors predict better but inform a decision that has already been made for most patients. The model is temporally (internally) validated but not yet externally validated, and because it was developed on a single platform it should be regarded as a recalibration target rather than a drop-in deployment elsewhere. The tool is published as a public web calculator to support shared decision-making, though it is not precise enough on its own to drive an irreversible clinical decision. It is prognostic, not therapeutic; treatment-effect estimation is addressed in companion work.

## 1. Introduction

Most published GLP-1 prediction models study only the patients who finish six months. But most patients never get there: in real-world telehealth, the majority disengage before the six-month mark. A model trained on completers therefore answers a question about the wrong population, and it answers it at a point when the clinical decision has already passed. This tool does the opposite. It includes the patients who disengage, and it predicts at the week-8 visit, where the decision actually gets made.

GLP-1 medications have transformed obesity care. In randomized trials, semaglutide produces 14.9% weight loss over 68 weeks^1^ and tirzepatide produces 20.9% over 72 weeks at the highest dose.^2^ Telehealth has scaled access dramatically. Two facts make real-world decisions hard: patients respond very differently from one another, and most drop out before six months. At the week-8 visit, a clinician needs three things at once: a calibrated prediction of where the patient will land at six months, the probability the patient drops out before then, and a realistic timeline for plateau. We supply all three.

This is a prognostic tool, not a treatment recommendation. It says where the patient is headed if current care continues; it does not say what to change. We target the accuracy needed to calibrate expectations and identify candidates for further clinical assessment, not the accuracy needed to drive an irreversible decision on its own. We make this distinction explicit in Section 3.2 and in the clinical-utility framing in Section 4.4. Estimating treatment effects is a separate problem requiring causal methods to address titration phase, confounding by indication, and unobserved engagement; that work appears in our companion paper.

## 2. Methods

### 2.1 Cohort

The full cohort comprised 237,800 adults enrolled in the Mochi Health telehealth GLP-1 program with a documented baseline weight between January 2020 and April 2026; cohort flow appears in Figure 1. Of these, 185,425 had at least one weight observation after starting therapy. Requiring a documented week-8 weight and a refill-confirmed dose left 68,969 patients, and further requiring reported ethnicity left an analytic cohort of 22,538. Within that cohort, 7,693 patients had a documented six-month outcome and form the Q1 prediction cohort; the full 22,538 drive the Q2 disengagement model; and the Q3 plateau cohort is 11,161 engaged patients with at least four weight observations.

**Figure 1:**
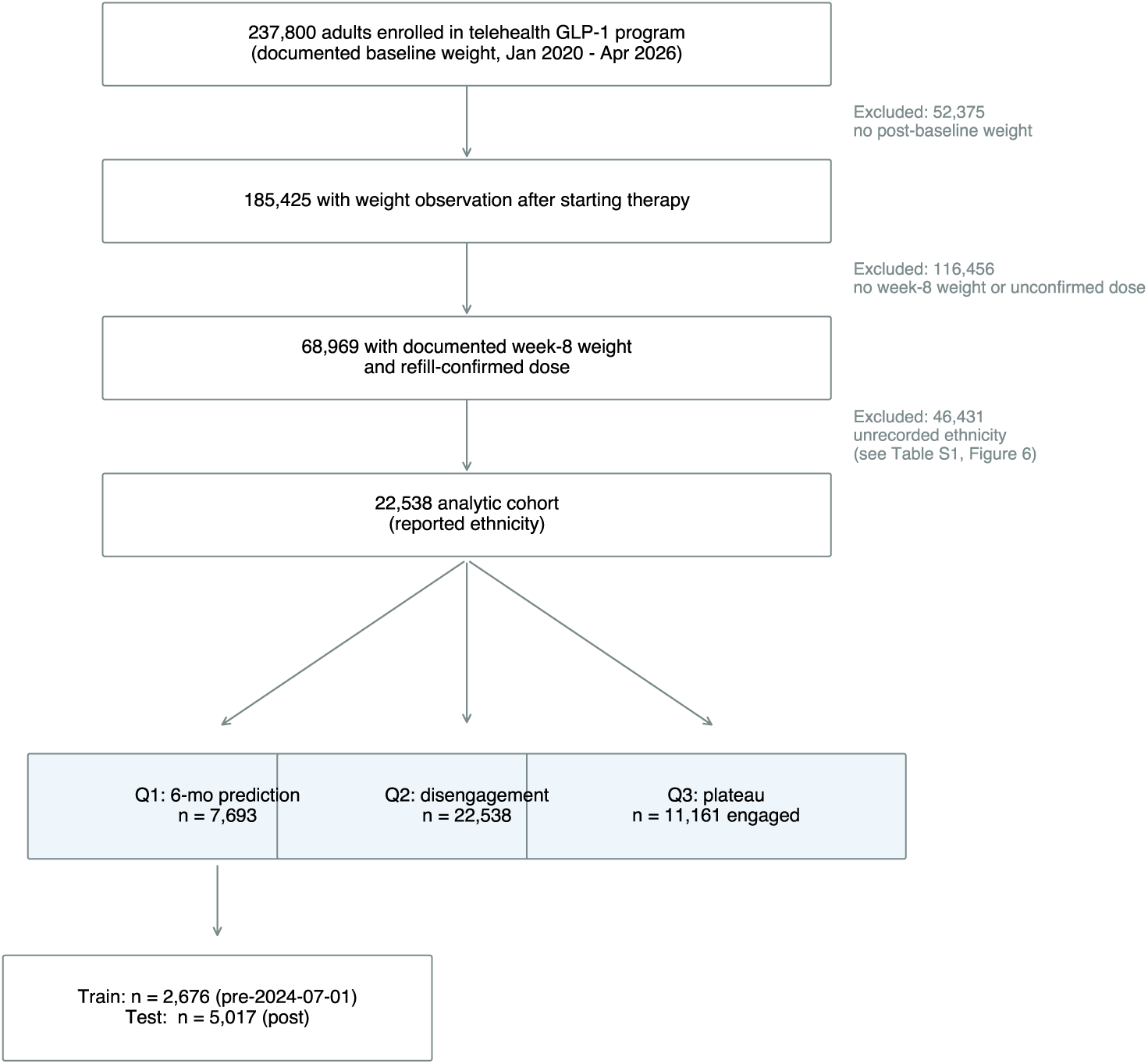
Cohort flow. The 237,800-patient master cohort is reduced to the 22,538-patient analytic cohort through three exclusion steps. The Q1 prediction cohort is split temporally for training and testing. The 46,431 patients excluded for unrecorded ethnicity are not exchangeable with the included cohort; see Figure 4 and Table S1.

### 2.2 Operational definitions

Three terms carry analytic weight, and we define each explicitly.

**Six-month disengagement** is the absence of any weight observation in days 140 to 210 from the patient’s program start. This is an operational endpoint, not a formal program-leaving definition, and it conflates several states: the patient who left the platform, the patient who is still enrolled but stopped self-reporting weights, and the patient who lost or replaced their scale. The data cannot distinguish these. The Q2 model therefore predicts the union of all of them: any reason the platform stops receiving a weight by month six. Throughout, “disengagement” means this operational endpoint.

**Slow responder** is a patient who has been on a therapeutic GLP-1 dose (Whitley tier 3 or higher: semaglutide *≥* 1.0 mg per week or tirzepatide *≥* 5 mg per week)^3^ for at least four weeks and has lost less than 5% of baseline weight at the time of measurement (within *±*14 days). The 5% threshold is consistent with FDA labeling and AACE/ACE clinical practice guidelines.^7^ A slow responder has reached and tolerated a therapeutic dose; we do not apply the label to patients still in titration.

**Post-titration index date** is the patient-specific day on which a patient first satisfies the slow-responder definition: the day of the first refill at therapeutic dose plus 28 days, conditional on at least one additional same-or-higher-tier refill in days 21 to 60 thereafter. This date marks when the post-titration decision becomes available to a clinician. Earlier dates are still in titration; later dates have already passed the decision window. We use the post-titration index date in two ways: as the index time for the causal analysis in the companion paper, and as the empirical reference point for assessing the actionability of the week-8 prediction (Section 2.7).

### 2.3 Outcomes

The primary outcome was percent weight loss from baseline at six months, taken from the most recent weight in days 140 to 210. Secondary outcomes were six-month disengagement, defined above, and time to reach 95% of an asymptote-fitted plateau among engaged patients.

### 2.4 Q1: six-month outcome prediction

We fit a cubic in observed week-8 percent loss plus 16 covariates: baseline BMI, weight, age, sex, drug class at week 8, week-8 dose, side-effect counts, severity, GI flag, goal-percent target, dietitian visit, exercise minutes per week, and four ethnicity buckets (Other is the reference). We built prediction intervals two ways: with a constant residual standard deviation, and with quantile regression at the 10th, 90th, 2.5th, and 97.5th percentiles. We compared a fractional-logit version of the model against ordinary least squares and, where sample size allowed, recalibrated each subgroup with isotonic regression.

We split by enrollment date, training on enrollments before 2024-07-01 (n = 2,676) and testing on later ones (n = 5,017). This temporal validation split is the design recommended by TRIPOD+AI^8^ and PROBAST^11^ for prognostic models intended for prospective use: the model is trained on past patients and tested on a subsequent cohort, so the reported test performance reflects how the model will behave on the next patient through the door. The fold sizes are inverted (test larger than train) because the platform’s enrollment grew rapidly across the split date. To rule out that the smaller training fold drives the headline R^2^, we report two sensitivity analyses (Section 3.6): a reversed-direction split (train on the post-split fold, test on the pre-split fold) and a random 80/20 split. We assessed calibration by binning test predictions into deciles, and we report calibration-in-the-large and slope on the full test set.

### 2.5 Per-group recalibration and subgroup flags

Because under-prediction can differ across subgroups, we recalibrated each subgroup with isotonic regression on a 50-50 split of the test set. The recalibration mapping was fit on test fold 0 (n = 2,509) and evaluated on test fold 1 (n = 2,508), which was untouched by any earlier model fitting or hyperparameter choice. We recalibrated only subgroups with at least 50 patients in the fit fold. If a subgroup had fewer than 30 evaluation-fold patients, the calculator suppresses a prediction; if subgroup MAE was more than 0.30 pp worse than the cohort mean, the calculator shows a reduced-accuracy disclosure.

### 2.6 Q2: disengagement prediction

We fit a logistic regression of binary disengagement at six months on the same 16 covariates plus week-8 percent loss, benchmarked against gradient boosting (200 trees, max depth 4). Performance was measured by area under the receiver operating characteristic curve on temporal hold-out.

### 2.7 Q3: plateau timing

We fit each patient’s trajectory to *y*(*t*) = *L*(1 *− e^−kt^*), where *L* is the asymptotic percent loss and *k* is the rate constant, restricting fits to patients with at least four weight observations (n = 11,161). For each patient we computed the day at which the curve reaches 95% of its asymptote, and we stratified by drug class at week 8 and by baseline BMI tertile. Because this restriction selects for engaged patients, the plateau time we report applies to engaged patients only. Population-level plateau estimates would require modeling the trajectory and informative censoring jointly, which we leave to future work.

### 2.8 Anchor choice as a trade-off

The choice of measurement-week anchor balances three properties: titration completion, predictive accuracy, and actionability.

#### Titration completion

The FDA labels for both drugs require a titration schedule lasting 12 to 20 weeks. Anchoring earlier than week 8 means most patients are still on sub-therapeutic doses, so the prediction rests on a weight-loss signal generated by the protocol itself rather than by the patient’s biology.

#### Predictive accuracy

R^2^ is generally increasing (with a small non-monotone dip at weeks 10-12 due to differing per-anchor subsets) with the anchor week: earlier weeks predict less well, later weeks better.

#### Actionability

The clinical decision to escalate, switch, or continue is meaningful only as long as it is still on the table. Once the post-titration index date passes, the decision has already been made for that patient, whether actively or by default.

We chose week 8 because it is the earliest visit at which all three properties hold simultaneously: the majority of patients have completed titration, R^2^ has reached a clinically useful range, and the post-titration decision window is still open. Later anchors (weeks 12, 16, 20) predict more accurately but inform a decision already made for most patients. We quantify each property below by refitting the cubic outcome model at weeks 2, 4, 6, 8, 10, 12, 16, and 20 and reporting test R^2^ and MAE, and by reporting the distribution of post-titration index dates among confirmed slow responders.

### 2.9 Decision curve analysis

We computed net benefit across the computed threshold range of 0.05 to 0.50 for the binary decision “predict at-risk for poor outcome,” comparing the model to “treat all” and “treat none” strategies following Vickers and Elkin.^6^

### 2.10 Benchmark calibration

We computed mean six-month weight loss in completers, separately for semaglutide and tirzepatide, and compared it to the six-month outcomes reported in STEP-1^1^ and SURMOUNT-1.^2^

### 2.11 Software

We used Python 3.11 with scikit-learn 1.4, pandas 2.1, statsmodels, scipy, and matplotlib. Quantile regression was fit using scikit-learn’s QuantileRegressor with the HiGHS solver.

## 3. Results

### 3.1 Cohort and benchmark calibration

The analytic cohort comprised 22,538 patients (Table 1). Mean age was 43.7 years, 92% of patients were female, and mean baseline BMI was 30.4 kg/m^2^. At week 8, 56% were on tirzepatide and 44% on semaglutide. Mean six-month weight loss in completers was 11.7% on semaglutide and 14.1% on tirzepatide. Six-month disengagement was 66%.

**Table 1.** Cohort characteristics. The analytic cohort and the Q1 prediction cohort. Continuous variables shown as mean (SD); categorical as n (%).

|  | Analytic (n = 22,538) | Q1 prediction (n = 7,693) |
| --- | --- | --- |
| Age, mean (SD) | 43.7 (11.8) | 43.4 (11.2) |
| Female, n (%) | 20,693 (92%) | 7,121 (93%) |
| BMI, mean (SD) | 30.4 (6.9) | 29.4 (6.5) |
| Weight (lb), mean (SD) | 206.7 (51.4) | 218.0 (50.4) |
| Tirzepatide at w8, n (%) | 12,543 (56%) | 4,115 (53%) |
| Goal % loss, mean (SD) | 23.0 (10.9) | 27.0 (9.8) |
| Met dietitian, n (%) | 9,955 (44%) | 4,091 (53%) |
| Eth: White, n (%) | 15,661 (69%) | 5,578 (73%) |
| Eth: Hispanic, n (%) | 2,605 (12%) | 801 (10%) |
| Eth: Black, n (%) | 1,586 (7%) | 472 (6%) |
| Eth: Asian, n (%) | 877 (4%) | 296 (4%) |
| Eth: Other, n (%) | 1,809 (8%) | 546 (7%) |

### 3.2 Q1: six-month outcome prediction

The cubic model on week-8 percent loss plus 16 covariates achieved training R^2^ = 0.71 and test R^2^ = 0.65, with MAE 2.76 pp on n = 5,017 (Table 2). Five-fold cross-validation on the training fold gave R^2^ = 0.69 (SD 0.04).

**Table 2.**
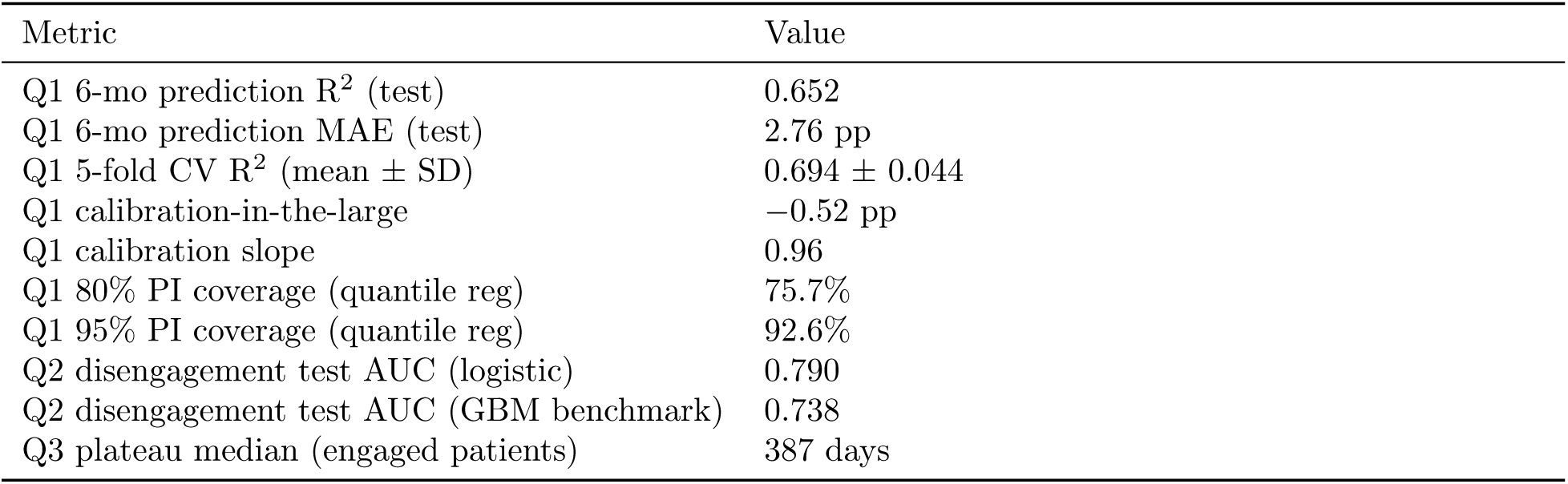
Prediction model performance summary.

These numbers have a clear clinical reading. R^2^ = 0.65 means the model explains 65% of the variance in six-month percent loss, leaving 35% of patient-to-patient variation unexplained by week-8 weight and the 16 covariates. MAE = 2.76 pp on a mean six-month loss of approximately 12% means a typical prediction is off by about 20 to 25% of the outcome’s magnitude. By the conventions of the prognostic-modeling literature,^8,9^ this level of accuracy is appropriate for a tool meant to calibrate expectations and identify candidates for further clinical assessment; it is not precise enough on its own to drive an irreversible clinical decision such as drug change, dose escalation, or surgical referral. The prediction interval (Section 3.4), not the point estimate, is the operative guide for clinical use.

Week-8 percent loss alone already carries most of the signal: a linear fit on week-8 loss gives R^2^ = 0.55, and adding the cubic term plus the 16 covariates beyond week-8 raises this by about 0.06 to R^2^ = 0.65. In other words, observed week-8 weight loss largely carries forward to the six-month outcome, and the covariates add modestly on top of it. One caveat tempers even this modest gain: the predictor (week-8 percent loss) and the target (six-month percent loss) share the same baseline-weight denominator and the same self-reported weighing instrument, so part of their apparent correlation reflects construct overlap rather than independent predictive content.

The calibration plot (Figure 2) shows residual error of approximately *−*0.5 percentage points across the lower deciles, with the top decile slightly under-predicted by 1.5 percentage points. Calibration-in-the-large was *−*0.52 pp and the calibration slope was 0.96 (perfect = 1.0).

**Figure 2:**
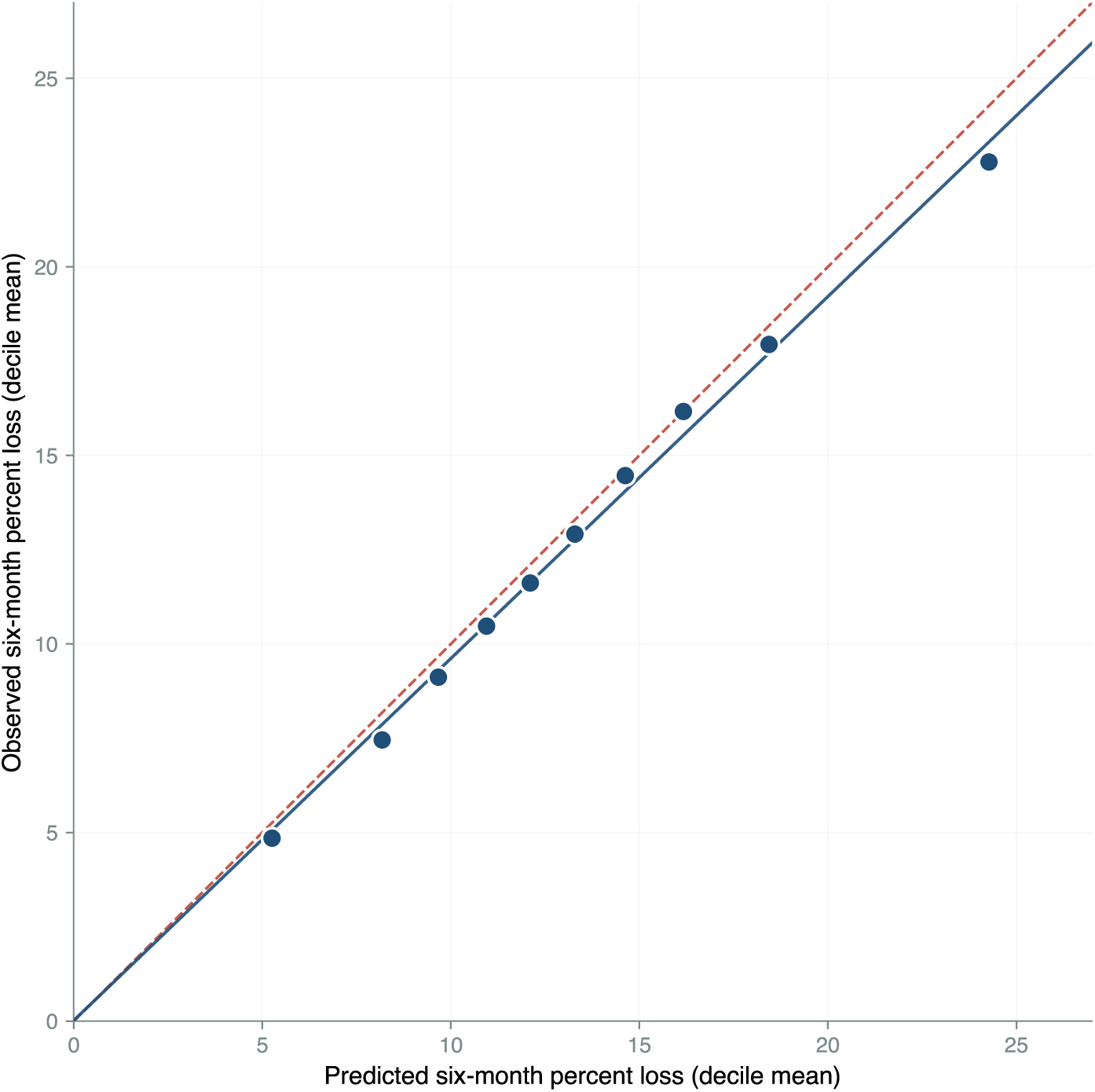
Calibration of the six-month prediction model. Predicted versus observed mean six-month percent loss by decile of model prediction (n = 5,017 test patients). The dashed reference line is identity (perfect calibration). Test R^2^ = 0.65, MAE = 2.76 pp, calibration-in-the-large = *−*0.52 pp, slope = 0.96. Both calibration metrics fall within published norms for prognostic models.^8,9^

Figure 3 shows per-patient prediction accuracy across all 5,017 test patients. The mass of patients falls along the identity line, and the shaded band shows the 80% prediction interval. The behavior covariates (dietitian visit, exercise minutes) contribute essentially nothing to the six-month prediction once observed week-8 loss is accounted for: their marginal R^2^ contribution is approximately zero, and the partial coefficients are *−*0.24 pp for dietitian visit (p = 0.05) and roughly zero for exercise (p = 0.33). Both behaviors are stronger predictors of week-8 loss itself (dietitian: *β* = *−*0.23, p *<* 10*^−^*^4^; exercise: *β* = +0.0014/min, p = 0.002), so the model captures behavior through the week-8 anchor rather than through the behavior covariates directly. The behavior gap over time is shown in Supplementary Figure S5.

**Figure 3:**
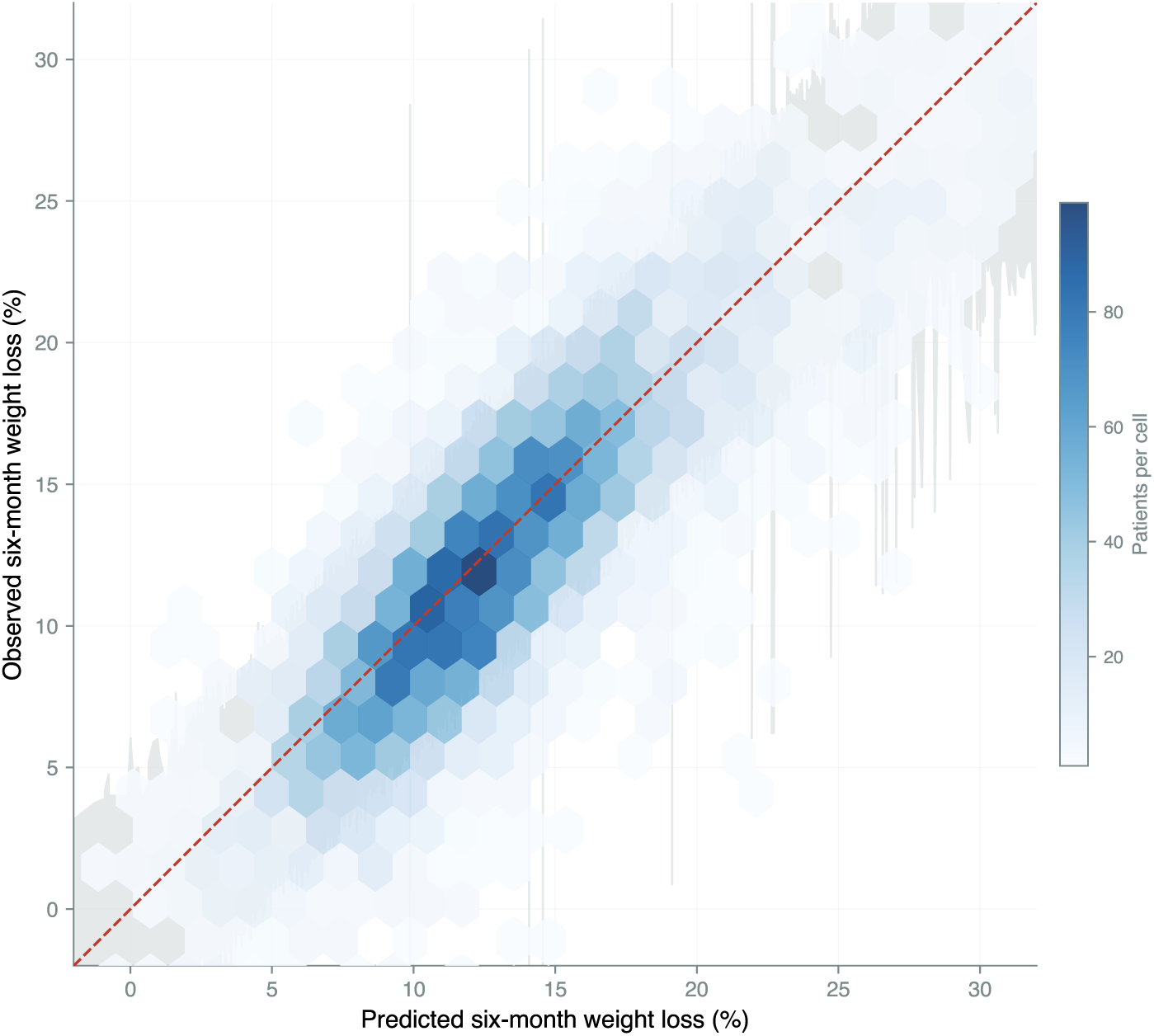
Per-patient prediction accuracy. Density-binned scatter of predicted versus observed six-month percent loss across all 5,017 test patients. Color intensity is the number of patients per cell. The dashed reference line is identity (perfect prediction). The shaded band is the 80% quantile-regression prediction interval.

Quantile-regression prediction intervals achieved 76% empirical coverage at 80% nominal and 93% at 95% nominal. Constant-residual-SD intervals under-covered (75% and 91%, respectively). Both quantile-regression intervals are slightly under-covered relative to nominal, by 4 percentage points at 80% and 2 percentage points at 95%, so the reported intervals are realistic but mildly optimistic and a clinician should treat them as approximate rather than guaranteed. A fractional-logit model gave essentially the same calibration as ordinary least squares; the under-prediction signature is structural rather than a link-function artifact (Supplementary Figure S4).

### 3.3 Sensitivity to the temporal split

The primary split trains on enrollments before 2024-07-01 (n = 2,676) and tests on later ones (n = 5,017). Two sensitivity analyses confirm the result. The reversed-direction split (train n = 5,017, test n = 2,676) gave test R^2^ = 0.658, MAE = 2.57 pp. A random 80/20 split (train n = 6,154, test n = 1,539) gave test R^2^ = 0.706, MAE = 2.54 pp. The headline R^2^ of 0.65 holds across all three splits, with the random split slightly better as expected when train and test are drawn from the same era. The inverted fold sizes in the primary split are a consequence of platform growth, not a methodological choice, and they do not drive the result.

### 3.4 Patients excluded from the analytic cohort

Of 68,969 patients with a documented week-8 weight and refill-confirmed dose, 22,538 (33%) had reported ethnicity and form the analytic cohort; 46,431 (67%) had unrecorded ethnicity and were excluded. The two groups differ on several baseline characteristics (Figure 4; full table in Supplementary Table S1). Standardized mean differences exceed 0.1 in absolute value for baseline BMI, exercise minutes per week, dietitian engagement, baseline weight, male sex, drug class at week 8, and goal percent loss. The included cohort has lower BMI, more female patients, and higher behavioral engagement than the excluded pool. Six-month disengagement is 79% in the excluded group versus 66% in the included (analytic) group. We discuss the implications for external validity in Section 4.5.

**Figure 4:**
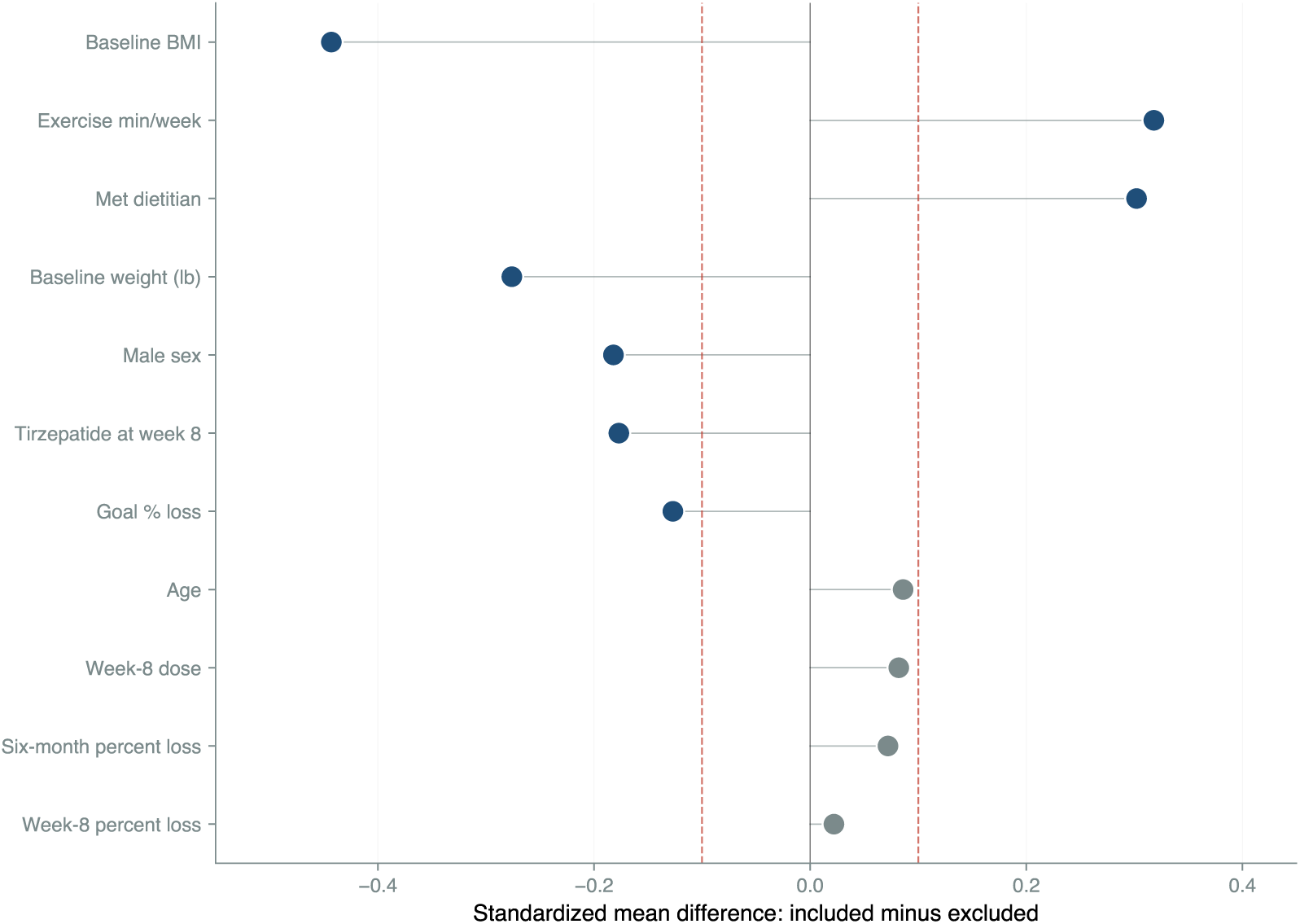
Selection bias on baseline characteristics. Standardized mean differences between the included cohort (n = 22,538, reported ethnicity) and the excluded cohort (n = 46,431, unrecorded ethnicity) for variables available in both groups. Vertical dashed lines mark the conventional *±*0.1 reference. Variables with *|*SMD*|* > 0.1 are highlighted; variables within *±*0.1 are shown in neutral. The included cohort is lower-BMI, lower-weight, more female, more often on semaglutide, and behaviorally more engaged than the excluded pool.

### 3.5 Empirical evidence on the anchor trade-off

The conceptual argument for the week-8 anchor is in Section 2.8; here we report the empirical inputs to that argument.

Test R^2^ rises with later anchor weeks: 0.48 at week 2, 0.57 at week 4, 0.61 at week 6, 0.65 at week 8, 0.72 at week 10, 0.67 at week 12, 0.87 at week 16, and 0.91 at week 20 (Figure 5). The week 10 to week 12 dip is non-monotonic because each anchor uses a different subset (patients with weight at that week): n drops from 3,962 to 3,667 with a different patient mix between the two windows. The overall pattern is generally increasing (with a small non-monotone dip at weeks 10-12 due to differing per-anchor subsets) in lead time.

**Figure 5:**
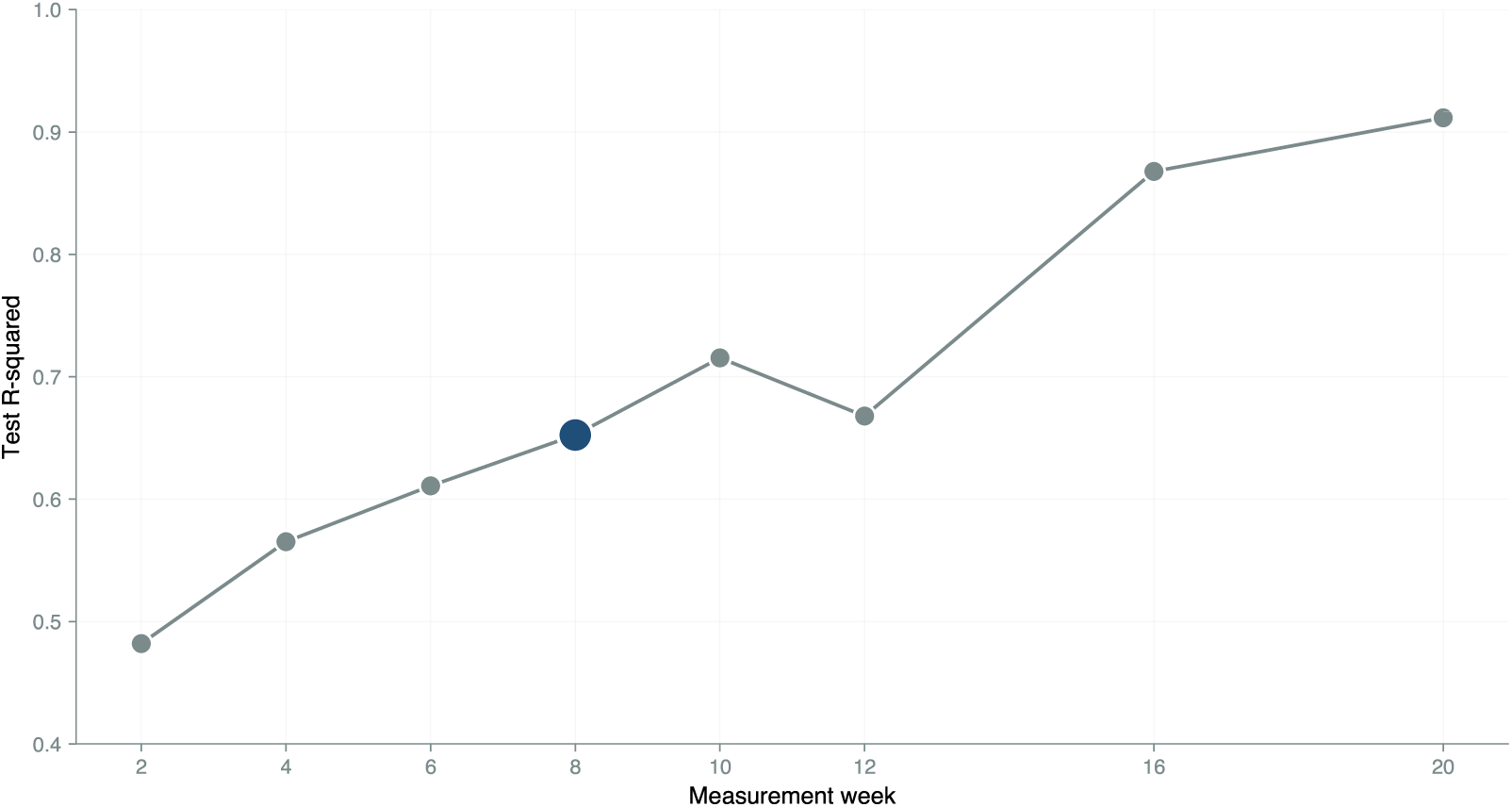
Predictive accuracy by measurement-week anchor. Test R^2^ at each measurement-week anchor. The week-8 anchor is highlighted in accent. R^2^ is generally increasing (with a small non-monotone dip at weeks 10-12 due to differing per-anchor subsets) with lead time. The conceptual trade-off motivating week 8 is described in Section 2.8.

Among 7,652 confirmed post-titration slow responders (a different denominator from the 7,693-patient Q1 outcome cohort, which is defined by a documented six-month outcome rather than by slow-responder status), the post-titration index falls at or before week 8 in 80% of cases (mean lead time *−*3.3 weeks, median *−*4.0; Figure 6). For these patients a prediction at week 8 is concurrent with the decision rather than anticipatory.

**Figure 6:**
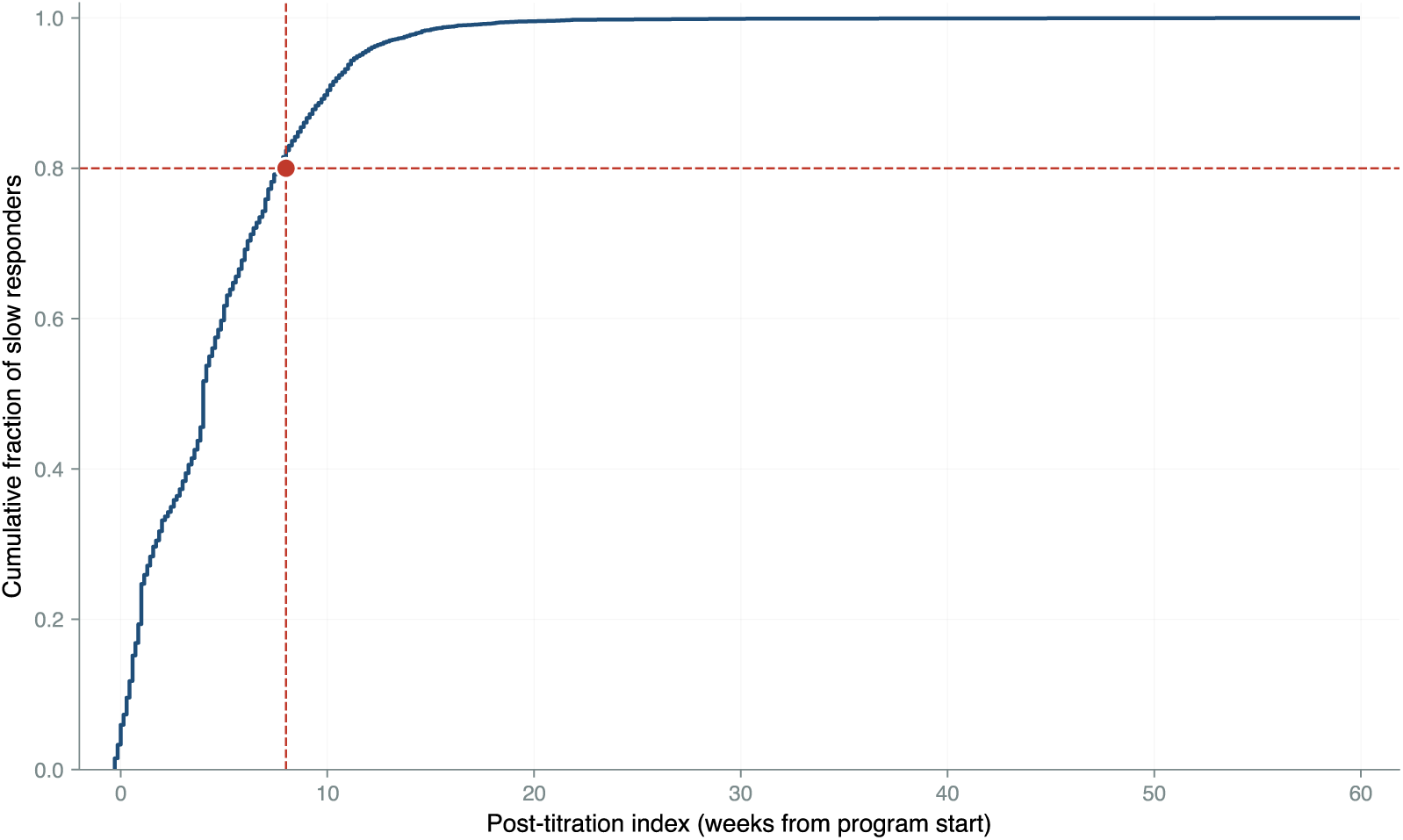
Post-titration index, cumulative distribution. Cumulative fraction of confirmed slow responders (n = 7,652) reaching the post-titration index by each week. The horizontal reference at 0.80 intersects the curve at week 8: 80% of slow responders reach the post-titration decision point at or before the week-8 visit.

### 3.6 Subgroup performance

Subgroup MAE in 20 ethnicity-by-age cells ranged from 2.17 to 3.38 pp (Supplementary Figure S1). Two cells were flagged for reduced-accuracy disclosure (Black 18-35, Asian 18-35), and two were too sparse to validate (Asian 52+ with n = 16, and n = 23 in another). Per-group recalibration improved MAE by 0.10 to 0.20 pp in the largest cells but added noise in smaller cells.

### 3.7 Q2: disengagement prediction

Logistic regression achieved training AUC 0.72 and test AUC 0.79 (Figure 7). Test AUC slightly exceeds training AUC because of the temporal split: later enrollments have higher disengagement rates that the model captures cleanly, and L2 regularization prevents over-fitting to the smaller training fold. Gradient boosting achieved test AUC 0.74, so the interpretable model dominated the more complex alternative. Subgroup AUC was stable: female 0.79, male 0.79; semaglutide 0.77, tirzepatide 0.81; non-White 0.77, White 0.80. The largest disengagement drivers were male sex (coefficient +0.62), Asian ethnicity (*−*0.36), prior dietitian visit (*−*0.29), and Hispanic ethnicity (*−*0.29).

**Figure 7:**
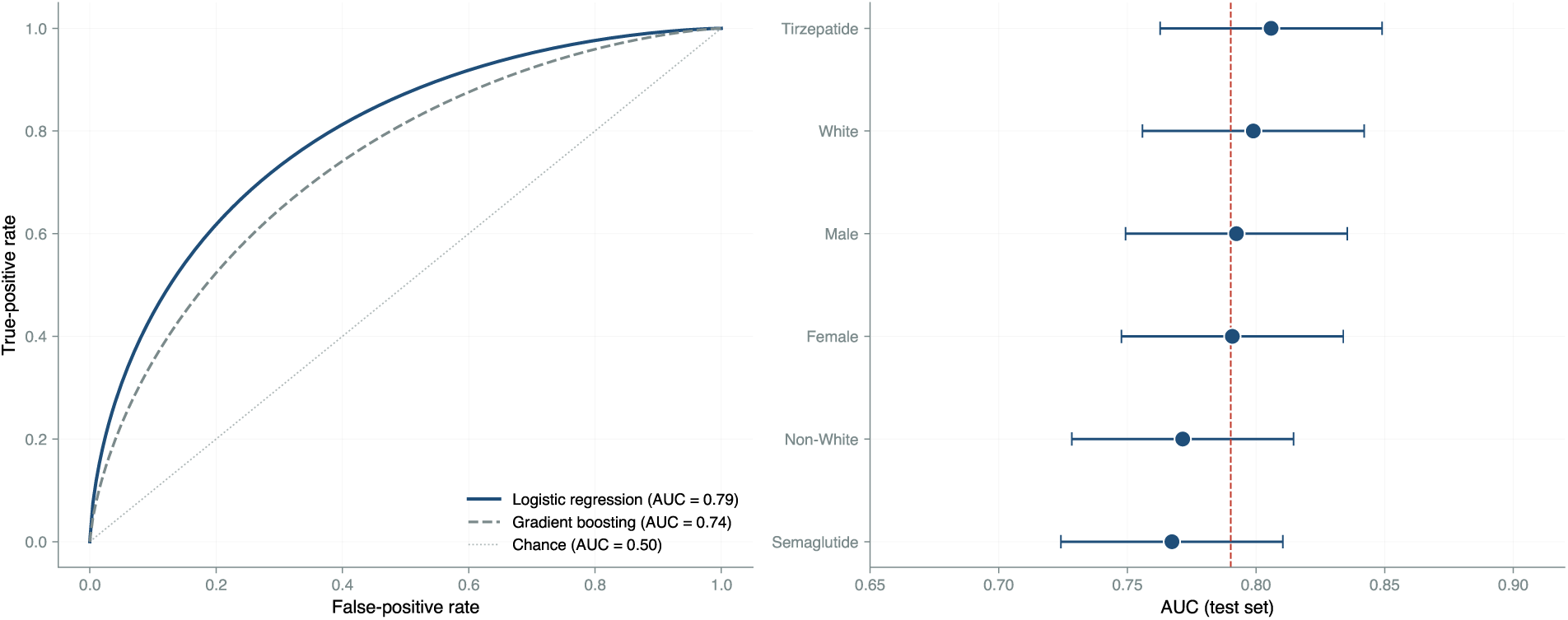
Disengagement model performance. (A) ROC curves for logistic regression and gradient boosting on temporal hold-out, with chance reference. AUCs are in the legend. (B) Subgroup AUC with 95% confidence intervals, sorted by point estimate. The dashed reference is the overall AUC. All subgroup point estimates lie within *±*0.03 of overall.

### 3.8 Q3: plateau timing

Across the 11,161 engaged patients, the median time to 95% of plateau was 387 days, with a 10th to 90th percentile range of 93 to 1,086 days and a median asymptote of 18.6%. This median is longer than the 327 days we estimated earlier on the unfiltered cohort: filtering for reported ethnicity selects a more engaged subpopulation, which plateaus later and at a higher loss.

By drug class, median plateau was 383 days for semaglutide and 389 days for tirzepatide. By baseline BMI tertile, the medians were 437 days (lowest), 394 (middle), and 321 (highest); the full distribution is shown in Figure 8. Lower BMI tracks with a later plateau and a higher asymptote.

**Figure 8:**
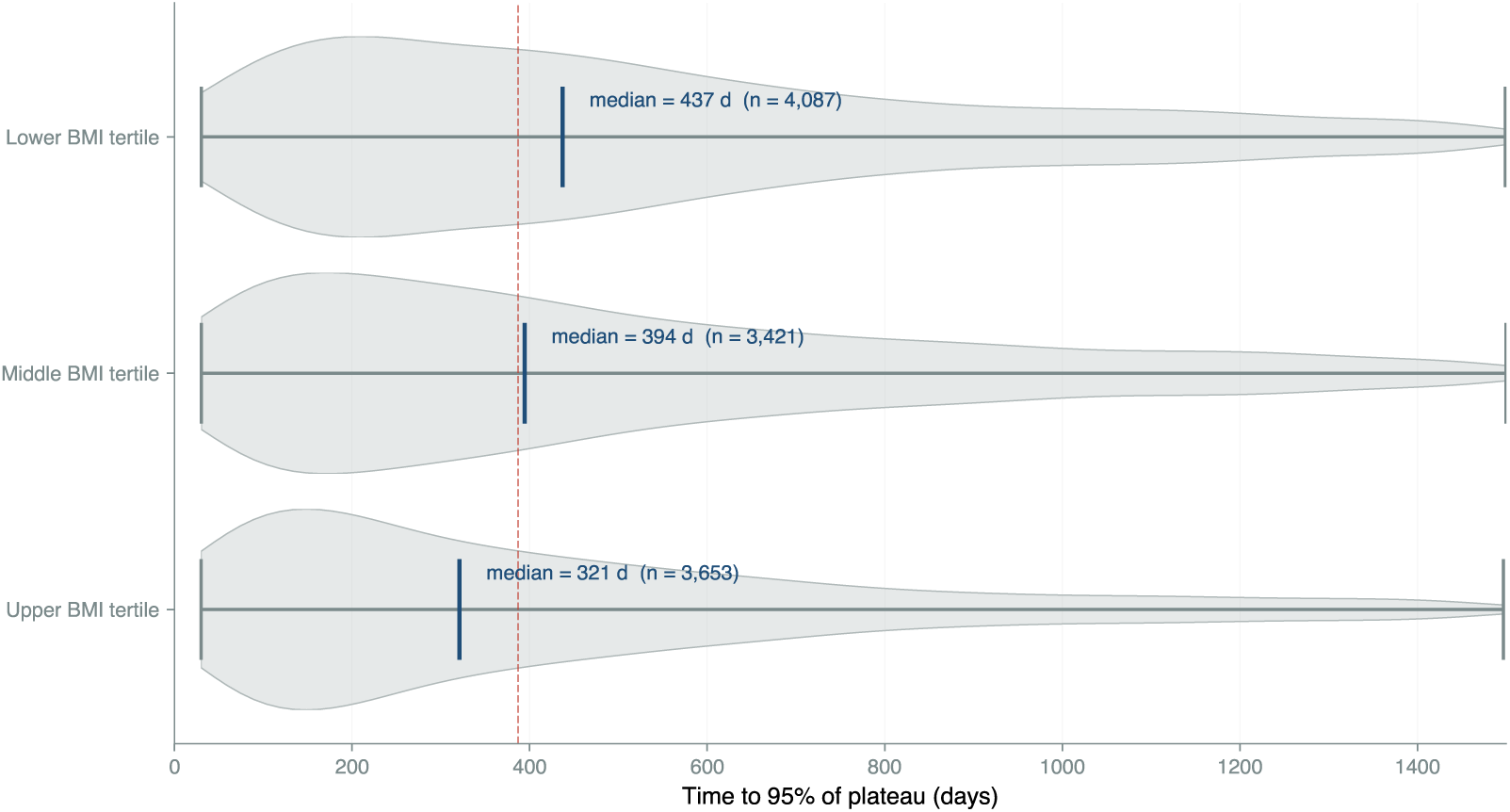
Plateau timing by baseline BMI tertile. Distribution of patient-specific time to 95% of asymptote among engaged patients (n = 11,161). The accent vertical bar in each violin marks the tertile median; the dashed reference at 437 days marks the lower-BMI median. Higher-BMI patients plateau substantially earlier and at a lower asymptote.

These estimates apply to engaged patients only. The full dropout-inclusive population has shorter follow-up on average, so a population-wide plateau would require joint modeling of trajectory and informative censoring.

### 3.9 Decision curve analysis

Over the computed threshold range of 0.05 to 0.50 (Section 2.9), the model achieved positive net benefit across the positive-net-benefit range of 0.05 to 0.45 (Figure 9), dominating “treat all” beyond threshold probability 0.10 and “treat none” across the entire range. We anchor the clinically-relevant threshold range at 0.10 to 0.30 because the prevalence of poor outcome (predicted six-month loss *<* 8%) in the test cohort is 22.3%; following the Vickers-van Calster-Steyerberg recommendation that the threshold range bracket the outcome prevalence,^10^ this range corresponds to clinicians willing to flag a patient as at-risk when their probability of poor outcome is roughly half to one-and-a-half times the population baseline. Outside this range the threshold is either so loose that it flags everyone or so strict that the flagged action is meaningless to apply.

**Figure 9:**
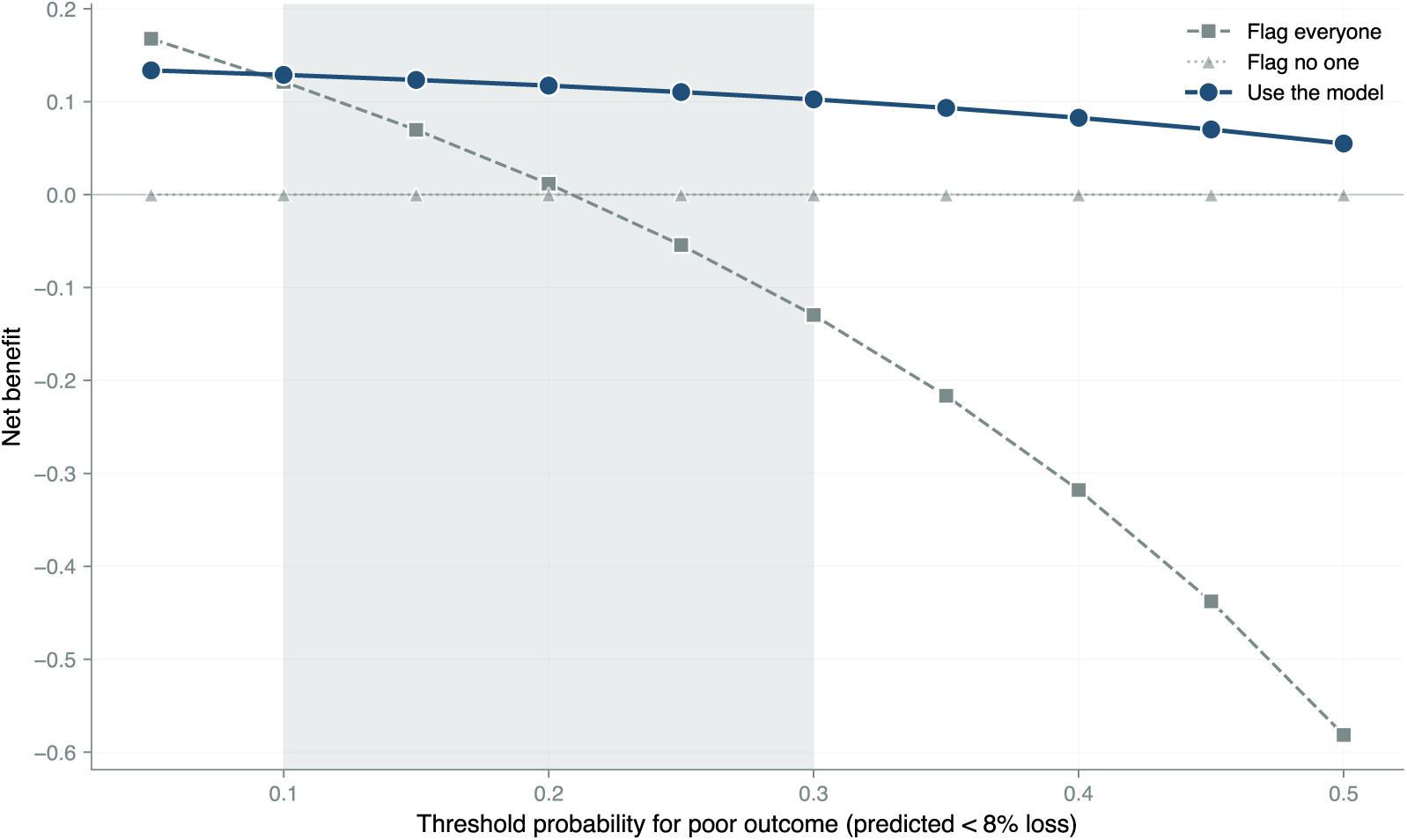
Decision curve analysis.^6,10^. Net benefit at each threshold probability for calling a patient at-risk (predicted *<*8% loss at six months). The shaded band is the threshold range bracketing the test-cohort poor-outcome prevalence (22.3%). The model achieves higher net benefit than flag-everyone and flag-no-one strategies across this range.

### 3.10 Decision-support calculator

The integrated tool implements the temporally (internally) validated, not yet externally validated cubic model with quantile-regression PIs, the disengagement logistic, and the plateau distribution. Required inputs are starting weight, week-8 weight, and goal weight in pounds; optional inputs refine the prediction. The tool returns predicted weight at six months with a calibrated 95% PI, probability of reaching the patient’s stated goal, disengagement probability, and predicted plateau day. It is prognostic and does not recommend treatment changes.

## 4. Discussion

### 4.1 Principal findings

We report three findings. First, observed week-8 weight loss is the dominant predictor of six-month outcome (test R^2^ = 0.65). Second, six-month disengagement is predictable at week 8 with AUC 0.79, and a logistic model dominates a tree ensemble. Third, plateau timing among engaged patients has a median of 387 days with substantial heterogeneity by drug class and baseline BMI.

### 4.2 The week-8 anchor is decision-concurrent

Week 8 is when most slow-responder decisions are actually made. Eighty percent of confirmed slow responders reach the post-titration index at or before week 8, so a prediction at that visit informs the live decision rather than a future one. Later anchors predict better, but by then the decision has already been made for most patients. Week 8 is also the platform’s standard check-in cadence, so anchoring the prediction there matches how the visit is already structured.

### 4.3 Limitations

Five limitations bear on interpretation.

#### Subgroup accuracy

Predictive accuracy is reduced for young Black and Asian patients (MAE about 0.4 pp worse than the cohort average). The calculator discloses this, and we are working on a calibration update with a larger minority sample.

#### Sparse cells

Four subgroup cells had too few patients to validate. The calculator suppresses predictions for those cells rather than predicting and disclaiming.

#### Selection on ethnicity reporting

The analytic cohort excludes 46,431 patients (67% of the master cohort) whose ethnicity is unrecorded, and the two groups are not exchangeable. Excluded patients have higher BMI by 3.1 kg/m^2^, are 6 percentage points more often male, are 8 percentage points more often on tirzepatide, and have 14 percentage points lower dietitian engagement (Figure 4; full numerical table in Supplementary Table S1). Six-month disengagement is 79% in the excluded group versus 66% in the included group. Because the tool’s calibration was assessed on the included subpopulation, we cannot assert the same calibration in the broader pool of patients without reported ethnicity. Two practical implications follow. First, the predicted six-month outcomes will likely under-state the disengagement risk and may under-predict outcome variance for higher-BMI, less-engaged patients than the analytic cohort represents. Second, deploying the tool to a clinic whose patient mix resembles the excluded group (heavier, more male, less behaviorally engaged) is a recalibration target rather than a drop-in deployment. We recommend a per-clinic calibration check on the first 100 to 200 patients before relying on the prediction interval for shared decision-making.

#### Plateau is conditional on engagement

The 387-day median applies to the 11,161 patients (49% of the analytic cohort) with at least four weight observations; a population-level plateau would require joint modeling of trajectory and informative censoring.

#### Single platform, retrospective

This is a single-platform retrospective analysis, and external validation in independent telehealth cohorts is needed.

### 4.4 What the clinician does with the output

A common and reasonable critique of prognostic tools is that they report numbers without saying what the clinician should do with them. We address this directly. The tool produces three outputs at the week-8 visit: a predicted six-month percent loss with an 80% prediction interval, a probability of disengagement before six months, and a predicted plateau day among patients who stay engaged. The intended uses follow, with concrete vignettes from the model.

#### 1. Calibrate expectations and refine the goal

Most patients enter the program with a goal weight that may or may not be realistic on their current trajectory, and the tool’s primary use is to ground the conversation in a calibrated expectation. Consider a patient at week 8 with 5.5% loss, on tirzepatide, BMI 32, no dietitian visit: the predicted six-month outcome is 12.4% (80% PI 8.4 to 16.7%) with a 62% probability of disengagement. Their stated goal of 25% loss is outside the prediction interval; a realistic six-month expectation is the 8 to 17% range, with the lower end more likely if engagement drops.

#### 2. Identify candidates for the post-titration medication decision

Patients with low predicted outcomes at week 8 are the candidates for medication-side decisions: within-class escalation, equipotent class switch, or class switch with potency increase. Consider a patient at week 8 with 3% loss, on semaglutide, BMI 35, no dietitian visit: the predicted six-month outcome is 7.6% (80% PI 3.6 to 12.0%) with an 83% probability of disengagement. The point estimate is the lowest in our vignette set and the disengagement risk the highest. *This paper supports the identification step.* It does not by itself supply the evidence to choose among the medication-side options for that patient; the comparative effectiveness of each option in confirmed post-titration slow responders is the subject of the companion causal paper. The division is explicit: the prognostic model here tells the clinician who is below trajectory, while the causal model in the companion paper tells the clinician what each medication-side option is worth.

#### 3. Triage engagement support

High disengagement probability at week 8 is an actionable signal supported entirely by the present paper. Adding a dietitian visit to the same slow-responder vignette above (week-8 loss 3%, BMI 35) produces an essentially identical predicted outcome (7.7%) but a noticeably lower disengagement probability (79% vs 83%). The decision-curve analysis (Figure 9) shows that flagging a patient as at-risk based on the model is more useful than flagging everyone or no one across the threshold range a clinician would actually use. The action this evidence supports is engagement support (proactive outreach, scheduling a dietitian, reviewing tolerability), not a decision to alter the medication. Intensive behavioral and lifestyle support has independently demonstrated weight benefit in randomized trials of both pharmacologic and non-pharmacologic care.^4,5^

The tool does not recommend a specific drug, dose, or referral. It produces calibrated expectations, identifies who falls below those expectations, and identifies who is at risk of being lost to follow-up before the next decision visit. The medication-side decision is supported by the companion causal analysis; the engagement-triage decision is supported by the disengagement model presented here. That distinction matters for which claims this paper asks to be evaluated on.

## 5. Conclusions

In 22,538 patients on GLP-1 therapy through a US telehealth platform, observed week-8 weight loss predicts six-month outcome with R^2^ = 0.65 on temporal hold-out. Six-month disengagement is predictable with AUC 0.79. Engaged patients plateau at a median of 387 days, with meaningful variation by drug and BMI. Week 8 is the decision-concurrent anchor. The model is temporally (internally) validated but not yet externally validated, and because it was developed and calibrated on a single platform it should be treated as a recalibration target rather than a drop-in deployment in other settings. As a decision-support tool it is published as a web calculator and is intended to support shared decision-making at the week-8 visit, while remaining not precise enough on its own to drive an irreversible clinical decision. Treatment-effect estimation is addressed in companion work.

## Data Availability

Individual patient-level data are private and cannot be shared publicly. De-identified data, the analysis code, and the prognostic calculator can be provided upon reasonable request to the corresponding author.

## Author Contributions

**Brian Erly:** Conceptualization, Supervision, Writing, Clinical validation.

**Shanmugesh Raja:** Conceptualization, Data curation, Formal analysis, Methodology, Software, Visualization, Writing.

## Ethics Approval

This study is a secondary analysis of de-identified data collected during routine clinical care at Mochi Health. As the analysis involves no direct patient contact, no intervention, and uses only data deidentified per HIPAA Safe Harbor [45 CFR 164.514(b)(2)], it was determined to not constitute human subjects research as defined by 45 CFR 46.102. Formal IRB review was therefore not required.

## Reporting Guideline

This study is reported in accordance with the TRIPOD+AI statement;^8^ a completed TRIPOD+AI checklist is provided as a supplement [if available].

